# Population-based analysis of radiation-induced gliomas after cranial radiotherapy for childhood cancers

**DOI:** 10.1101/2022.03.04.22271880

**Authors:** Jacob B. Leary, Amy Anderson-Mellies, Adam L. Green

## Abstract

**Background:** Cranial radiotherapy (RT) is used to treat pediatric central nervous system (CNS) cancers and leukemias. RT carries a risk of secondary CNS malignancies, including radiation-induced gliomas, the epidemiology of which is poorly understood.

**Methods:** This retrospective study using SEER registry data (1975-2016) included two cohorts. Cohort 1 included patients diagnosed with Grade III/IV or ungraded glioma as a second malignancy at least 2 years after receiving beam radiation and/or chemotherapy for a first malignancy diagnosed at ages 0-19 years, either a primary CNS tumor treated with RT (1a, n=57) or leukemia with unknown RT treatment (1b, n=20). Cohort 2 included patients with possible missed RIG who received RT for a primary CNS tumor diagnosed at 0-19 and then died of presumed progressive disease more than 5 years after diagnosis, since previous studies have documented many missed RIGs in this group (n=296). Controls (n=10,687) included all other patients ages 0-19 who received RT for a first CNS tumor or leukemia who did not fit inclusion criteria above.

**Results:** For Cohort 1 (likely/definite RIGs), 0.97% of patients receiving cranial RT went on to develop RIG. 3.39% of patients receiving cranial RT for primary CNS tumors fell in Cohort 2 (potential RIGs). Median latency to RIG diagnosis was 11.1 years; latency was significantly shorter for Cohort 1b (median 10.0, range 5.0-16.1) vs. 1a (12.0, 3.6-34.4, p=0.018). Median OS for Cohort 1 was 9.0 months. Receiving surgery, radiation, or chemotherapy were all associated with a non-statistically significant improvement in OS (p 0.1-0.2). 1.8% of brain tumor deaths in the cohort fell in Cohort 1, with an additional 7.9% in Cohort 2.

**Conclusion:** Within the limitations of a population-based study, 1-4% of patients undergoing cranial RT for pediatric cancers later develop RIG, which is incurable and can occur anywhere from 3-35 years later. 2-10% of pediatric brain tumor deaths are attributable to RIG. Effective treatment of RIG remains unclear and is thus deserving of increased attention in preclinical and clinical studies.

## Introduction

Central nervous system (CNS) tumors are the 2^nd^ most common pediatric malignancy but the leading cause of cancer-related death in young patients. Of these, roughly 10% are classified as pediatric high-grade gliomas (pHGG), but these account for 40% of CNS tumor-related deaths^1^. Given the potentially devastating consequences of these malignancies in terms of significant early-life morbidity and mortality, it is crucial to better understand their behavior and to develop therapies aimed at reducing their negative impact on pediatric patients’ lives.

The current standard-of-care treatment for pHGG continues to be maximal safe resection followed by focal radiation therapy (RT). Resection provides greatest overall survival (OS) benefit regardless of other choices of treatment modalities^2,3^. However, pHGG is characterized by diffuse distribution with infiltrative growth patterns, often arising in difficult-to-reach, delicate, and vital portions of the brain, making complete surgical resection impossible^4,5^.

Fractionated external-beam radiation therapy (RT) is frequently used to target midline tumors, as well as against diffuse CNS tumors in other brain regions or to reduce tumor burden prior to attempting resection to increase the likelihood of success of surgical intervention.^6^ External-beam RT, which involves a radiation source of protons or photons located outside a patient’s body, is also commonly employed as an adjunct therapy to treat embryonal tumors such as medulloblastoma, as well as ependymoma and other CNS tumors.^7^ In pediatric leukemias, cranial RT has been used as preventive therapy for leptomeningeal spread and in cases of CNS disease at diagnosis; however, due to detrimental effects on cognition, as well as elevated risk of secondary malignancies, this has largely been replaced by intrathecal chemotherapy and/or significantly reduced doses of RT.^8^ Often, a combination of partial resection, RT, and chemotherapy is used to maximize the likelihood of total eradication or at minimum, to increase duration of progression-free survival and overall survival.^5,9^

RT exerts its effects via DNA damage, specifically through double strand breaks. The efficacy of this treatment is highly dependent on the amount of DNA damage induced.^10^ A more severe degree of DNA damage is more difficult for tumor cell DNA damage response (DDR) mechanisms to repair, increasing overall tumor cell death and thus more significantly decreasing tumor burden. However, DNA damage also occurs in healthy cells exposed to RT. Tremendous progress has been made over time to limit both dose and field of radiation exposure in order to minimize damage to healthy cells, which can lead to growth impairment, cognitive deficits, and secondary malignancies.^11^ Despite these strides in RT technique, some degree of inherent risk remains, especially when a larger tumor or larger portion of the brain requires irradiation.

Radiation-induced glioma/glioblastoma (RIG) is a high-grade secondary tumor arising in the CNS in regions previously irradiated, with or without systemic chemotherapy with potential for additional DNA damage, for a histologically distinct prior malignancy at an earlier age in childhood.^12^ These are thought to occur most commonly between 5-15 years following treatment for the primary malignancy and are poorly responsive to antitumor therapies including RT.^13^ RIG occurs most commonly following treatment for ependymoma, medulloblastoma, and leukemia.^14^ Prognosis for RIG is universally poor and often worse than other pHGG given that preclinical models have been difficult to develop, and no dedicated clinical trials have been conducted. As a result, treatment for RIG is far less standardized than for other subtypes of pHGG, and though resection similarly offers a longer duration of OS,^15^ outcomes remain largely grim. It is known that RIG often have a more homogeneous profile of mutations versus *de novo* pHGG, with more overlap and clustering of genetic signatures between RIG than in other pHGG. Whereas IDH and H3K27M mutations are common in other pHGG subtypes, they are extremely rare in RIG.^16,17^ Conversely, RIGs appear to cluster primarily with pHGG receptor tyrosine kinase 1 subtype based on DNA methylation, with copy-number alterations including Chromosome 1p loss/1q gain, and Chromosome 13q and 14q loss; they also frequently demonstrate other common mutations seen in the other pHGG subtypes including TP53, CDK4 amplification, CDKN2A/B and BCOR deletion, and amplification of receptor tyrosine kinases (BRAF, MET, and PDGFRA).^13,17,18^

Whereas molecular characteristics are beginning to be elucidated for RIG, little is known about the epidemiology of these secondary tumors. The purpose of the present study was to provide a characterization of patients with RIG derived from the Surveillance, Epidemiology. and End Results (SEER) Program registry, focusing on descriptive features of these tumors and the patients that they affect. Specifically, we aimed to define the true incidence of RIG including patterns of change in incidence over time, response to various therapeutic modalities, risk factors for RIG development, and the timeframe during which RIG development typically occurs following RT. We also sought to determine factors influencing overall survival (OS) of these tumors.

## Methods

### Study Design and Data Collection

This is a retrospective case-control study. Patient-level data were obtained from the SEER Program of the National Cancer Institute, a collection of population-based cancer registries throughout the United States. The years included in this study (1975-2016) encompass varying groupings of participating registries with population coverage ranging from 9% in 1975-1991 (SEER-9 registries) to 28% during 2000-2016 (SEER-18 registries). Patient demographics and cancer histories were abstracted from the SEER-18 Registries Custom Data, November 2018 Submission using SEER*Stat Version 8.3.6. Available information on patient age and tumor characteristics at time of diagnosis and first course of treatment was collected for each tumor in a patient’s history, as well as vital status, cause of death, and survival time.^19^

### Defining RIG

Constructing our cohort of patients diagnosed with RIG (Cohort 1) began with selecting those who had been diagnosed with a Grade III/IV or ungraded glioma as a second primary malignancy at least two years after receiving beam radiation and/or chemotherapy for a first primary malignancy diagnosed at age 0-19 years, and had a history of no more than two primary malignancies (n=124). The only patients included who had potentially undergone chemotherapy alone were patients with leukemia whose beam radiation treatment status was unknown; we elected to include these patients as possible RIG cases. Second primary glioma subtypes eligible for inclusion were as follows: anaplastic astrocytoma; diffuse astrocytoma; glioblastoma; oligodendroglioma; anaplastic oligodendroglioma; pilocytic astrocytoma; unique astrocytoma variants; mixed glioma; astrocytoma not otherwise specified (NOS); glioma NOS; benign and malignant neuronal/glial, neuronal, and mixed tumors; and unspecified CNS neoplasms. Patients with more than two primary malignancies were excluded, as this may be indicative of a tumor predisposition syndrome. Patient histories were then manually reviewed for primary tumor type/treatment and secondary tumor type. We excluded 44 patients whose first primary malignancy was neither CNS nor leukemia and patients whose second malignancy developed at a site presumed to be outside of the initial RT field. One patient with precursor T-cell lymphoblastic lymphoma affecting lymph nodes in multiple regions, and one patient with osteosarcoma of the mandible were excluded as their beam RT treatment status was no/unknown. We also excluded three additional patients with a first primary CNS malignancy whose beam radiation treatment status was no/unknown (n=1) or who received non-beam RT (n=2; one received radioactive implant brachytherapy and the other received RT, not otherwise specified). Cohort 1 was then further divided into those who were confirmed to have been treated with beam radiation for their first malignancy (Cohort 1a; n=57), and those patients with leukemia whose radiation treatment status was unknown for their first malignancy (Cohort 1b; n=20).

As a second cohort of possible undiagnosed RIG (Cohort 2), we included any other patient aged 0-19 who received beam radiation for a first primary CNS tumor whose death occurred 7 or more years after diagnosis (10 or more years for ependymomas, as these are known to have late true recurrences) and was attributed to their cancer. This cohort was included because it is now known that primary tumor recurrences this late after diagnosis are very rare, and many of these tumors may actually be RIGs that were either never biopsied or pathologically misclassified.^20,21^ This cohort was further divided into non-glioma (2a; n=139) and glioma (2b; n=157) as first malignancy to allow for distinction in the case that some of the Cohort 2b patients may have had a rare late recurrence rather than a RIG.

The control population for all cohorts included all other patients aged 0-19 who received beam radiation for a first primary CNS tumor or leukemia who did not fit the inclusion criteria for Cohorts 1 or 2 (Control; n=10,687).

### Outcomes of Interest

The primary outcomes of interest included demographic and tumor-specific characteristics for the RIG cohorts compared to controls, including age at initial diagnosis, sex, race, ethnicity, initial tumor type, and treatment type for first primary malignancy. Additionally, we sought to identify the incidence of RIG to characterize the overall risk of developing these tumors following treatment for the original pediatric malignancy. Other primary outcomes of interest included lag time between diagnosis of the first primary CNS tumor or leukemia and development of RIG, and median overall survival (OS) for patients who developed RIG, both measured in months.

Secondary outcomes of interest focused on incidence of RIG development broken down by treatment type for the first primary CNS tumor or leukemia. We also sought to evaluate median overall survival based on treatment type for RIG.

### Statistical Analysis

All statistical analyses were performed using SAS software, Version 9.4 (SAS Institute Inc., Cary, NC), with significance defined as p-value <0.05. All patients included in analyses had data available for all variables of interest in the SEER registries. The cohorts are used collectively to describe the occurrence of RIGs. Each cohort was compared to the control population on distributions of age at initial diagnosis, sex, race, ethnicity, initial tumor type, and treatment of initial tumor using Chi-square and Fisher’s exact tests. The proportion of CNS tumor deaths potentially attributable to RIGs was also estimated. Kaplan-Meier plots were used to visualize lag time between initial and RIG diagnoses, as well as to evaluate OS following RIG diagnosis. Univariate effects of treatment modality of RIG on survival were evaluated with the log-rank test.

## Results

Patient demographic and tumor-specific characteristics, including comparisons between groups, are displayed in Table 1. In terms of initial primary malignancies, Cohort 1a contained a predominance of medulloblastomas (38.6%), gliomas (26.3%), and leukemias (17.5%). Cohort 1b was composed entirely of leukemias treated with chemotherapy and with an unknown RT treatment status. Cohorts 2a and 2b were comprised of patients with secondary tumors that were considered possible RIGs, but were less clearly attributable to RT. In Cohort 2a, the most common original diagnoses were medulloblastoma (54.7%), PNET/pineal gland tumor (13.7%), germ cell tumor (13.0%), and ependymoma (12.2%). Cohort 2b was entirely composed of gliomas. See Table 1 for complete details for each group.

**Table 1.** Patient demographic and tumor-specific characteristics of RIG/possible RIG cohorts, compared to control cohort.

|  | Cohort 1a |  |  | Cohort 1b |  |  | Cohort 2a |  |  | Cohort 2b |  |  | Control |  |
| --- | --- | --- | --- | --- | --- | --- | --- | --- | --- | --- | --- | --- | --- | --- |
|  | N | % with attribute | P<sup>1</sup> | N | % with attribute | P<sup>1</sup> | N | % with attribute | P<sup>1</sup> | N | % with attribute | P<sup>1</sup> | N | % with attribute |
| **Overall** | 57 | 0.52 |  | 20 | 0.18 |  | 139 | 1.26 |  | 157 | 1.42 |  | 10,687 | 96.60 |
| **Age at Original Diagnosis** |  |  | 0.610 |  |  | 0.010 |  |  | 0.790 |  |  | 0.002 |  |  |
| <1 year | 1 | 1.75 |  | 1 | 5.00 |  | 1 | 0.72 |  | 7 | 4.46 |  | 213 | 1.99 |
| 1-4 years | 13 | 22.81 |  | 11 | 55.00 |  | 32 | 23.02 |  | 27 | 17.20 |  | 2,399 | 22.45 |
| 5-9 years | 21 | 36.84 |  | 4 | 20.00 |  | 44 | 31.65 |  | 40 | 25.48 |  | 3,113 | 29.13 |
| 10-14 years | 14 | 24.56 |  | 3 | 15.00 |  | 31 | 22.30 |  | 33 | 21.02 |  | 2,716 | 25.41 |
| 15-19 years | 8 | 14.04 |  | 1 | 5.00 |  | 31 | 22.30 |  | 50 | 31.85 |  | 2,246 | 21.02 |
| **Sex** |  |  | 0.091 |  |  | 0.706 |  |  | 0.633 |  |  | 0.205 |  |  |
| Female | 17 | 29.82 |  | 9 | 45.00 |  | 54 | 38.85 |  | 72 | 45.86 |  | 4,366 | 40.85 |
| Male | 40 | 70.18 |  | 11 | 55.00 |  | 85 | 61.15 |  | 85 | 54.14 |  | 6,321 | 59.15 |
| **Race** |  |  | 0.218 |  |  | 0.343 |  |  | 0.206 |  |  | 0.319 |  |  |
| White | 42 | 73.68 |  | 15 | 75.00 |  | 104 | 74.82 |  | 127 | 80.89 |  | 8,498 | 79.52 |
| Black | 5 | 8.77 |  | 1 | 5.00 |  | 23 | 16.55 |  | 19 | 12.10 |  | 1,149 | 10.75 |
| AI/AN | 0 | 0.00 |  | 0 | 0.00 |  | 2 | 1.44 |  | 3 | 1.91 |  | 96 | 0.90 |
| Asian/PI | 10 | 17.54 |  | 4 | 20.00 |  | 10 | 7.19 |  | 8 | 5.10 |  | 904 | 8.46 |
| Unknown | 0 | 0.00 |  | 0 | 0.00 |  | 0 | 0.00 |  | 0 | 0.00 |  | 40 | 0.37 |
| **Ethnicity** |  |  | 0.011 |  |  | 0.754 |  |  | 0.074 |  |  | <0.001 |  |  |
| Non-Hispanic | 52 | 91.23 |  | 16 | 80.00 |  | 116 | 83.45 |  | 145 | 92.36 |  | 8,235 | 77.06 |
| Hispanic | 5 | 8.77 |  | 4 | 20.00 |  | 23 | 16.55 |  | 12 | 7.64 |  | 2,452 | 22.94 |
| **Original Diagnosis** |  |  | <0.001 |  |  | -- |  |  | -- |  |  | -- |  |  |
| Glioma | 15 | 26.32 |  | 0 | 0.00 |  | 0 | 0.00 | -- | 157 | 100.00 | -- | 3,314 | 31.01 |
| Ependymoma | 3 | 5.26 |  | 0 | 0.00 |  | 17 | 12.23 | -- | 0 | 0.00 | -- | 821 | 7.68 |
| Choroid plexus tumor | 1 | 1.75 |  | 0 | 0.00 |  | 2 | 1.44 | -- | 0 | 0.00 | -- | 18 | 0.17 |
| Medulloblastoma | 22 | 38.60 |  | 0 | 0.00 |  | 76 | 54.68 | -- | 0 | 0.00 | -- | 1,624 | 15.20 |
| PNET/Pineal | 1 | 1.75 |  | 0 | 0.00 |  | 19 | 13.67 | -- | 0 | 0.00 | -- | 546 | 5.11 |
| ATRT | 1 | 1.75 |  | 0 | 0.00 |  | 1 | 0.72 | -- | 0 | 0.00 | -- | 112 | 1.05 |
| Germ cell tumor | 4 | 7.02 |  | 0 | 0.00 |  | 18 | 12.95 | -- | 0 | 0.00 | -- | 618 | 5.78 |
| Teratoma | 0 | 0.00 |  | 0 | 0.00 |  | 0 | 0.00 | -- | 0 | 0.00 | -- | 34 | 0.32 |
| Other/unspecified CNS | 0 | 0.00 |  | 0 | 0.00 |  | 6 | 4.32 | -- | 0 | 0.00 | -- | 183 | 1.71 |
| Leukemia | 10 | 17.54 |  | 20 | 100.00 |  | 0 | 0.00 | -- | 0 | 0.00 | -- | 3,417 | 31.97 |
| **Treatment of Original Tumor** |  |  | 0.089 |  |  | -- |  |  | 0.111 |  |  | <0.001 |  |  |
| Beam radiation, no/unknown chemotherapy | 22 | 38.60 |  | -- | -- |  | 48 | 34.53 |  | 122 | 77.71 |  | 3,035 | 28.40 |
| Beam radiation + chemotherapy | 35 | 61.40 |  | -- | -- |  | 91 | 65.47 |  | 35 | 22.29 |  | 7,652 | 71.60 |
| Chemotherapy, no/unknown beam radiation | -- | -- |  | 20 | 100.00 |  | -- | -- |  | -- | -- |  | -- | -- |
| **Histology of RIG** |  |  | -- |  |  | -- |  |  | -- |  |  | -- |  |  |
| Diffuse astrocytoma (protoplasma, fibrillary) | 1 | 1.75 |  | 0 | 0.00 |  | -- | -- |  | -- | -- |  | -- | -- |
| Anaplastic astrocytoma | 8 | 14.04 |  | 3 | 15.00 |  | -- | -- |  | -- | -- |  | -- | -- |
| Glioblastoma | 24 | 42.11 |  | 13 | 65.00 |  | -- | -- |  | -- | -- |  | -- | -- |
| Pilocytic astrocytoma | 0 | 0.00 |  | 0 | 0.00 |  | -- | -- |  | -- | -- |  | -- | -- |
| Oligodendroglioma | 1 | 1.75 |  | 0 | 0.00 |  | -- | -- |  | -- | -- |  | -- | -- |
| Anaplastic oligodendroglioma | 1 | 1.75 |  | 1 | 5.00 |  | -- | -- |  | -- | -- |  | -- | -- |
| Mixed glioma | 0 | 0.00 |  | 1 | 5.00 |  | -- | -- |  | -- | -- |  | -- | -- |
| Astrocytoma, NOS | 6 | 10.53 |  | 0 | 0.00 |  | -- | -- |  | -- | -- |  | -- | -- |
| Glioma, NOS | 13 | 22.81 |  | 1 | 5.00 |  | -- | -- |  | -- | -- |  | -- | -- |
| Benign and malignant neuronal/glial, neuronal and mixed | 1 | 1.75 |  | 1 | 5.00 |  | -- | -- |  | -- | -- |  | -- | -- |
| Neoplasm, unspecified, benign and malignant | 2 | 3.51 |  | 0 | 0.00 |  | -- | -- |  | -- | -- |  | -- | -- |

<sup>1</sup>P-value from Chi-square (or Fisher's Exact for cells <5) test comparing cohort to control

**Table 2.** Latency between original diagnosis and RIG diagnosis, in years.

|  | Mean | Std Dev | Median | Min | Max |
| --- | --- | --- | --- | --- | --- |
| Cohort 1 | 13.85 | 8.08 | 11.08 | 3.58 | 34.42 |
| Cohort 1a | 14.99 | 8.97 | 12.00 | 3.58 | 34.42 |
| Cohort 1b | 10.61 | 2.95 | 10.00 | 5.00 | 16.08 |

**Table 3.** Proportion of deaths from RIG overall and by original diagnosis.

|  | Cohort 1a |  | Cohort 1b |  | Cohort 2a |  | Cohort 2b |  | Total deaths<br>(Cohorts + Controls) |
| --- | --- | --- | --- | --- | --- | --- | --- | --- | --- |
|  | N | % of deaths | N | % of deaths | N | % of deaths | N | % of deaths |  |
| **Overall** | 50 | 1.00 | 16 | 0.32 | 139 | 2.79 | 157 | 3.15 | 4,988 |
| **Original Diagnosis** |  |  |  |  |  |  |  |  |  |
| Glioma | 13 | 0.55 | 0 | 0.00 | 0 | 0.00 | 157 | 6.69 | 2,347 |
| Ependymoma | 3 | 1.09 | 0 | 0.00 | 17 | 6.18 | 0 | 0.00 | 275 |
| Choroid plexus tumor | 1 | 10.00 | 0 | 0.00 | 2 | 20.00 | 0 | 0.00 | 10 |
| Medulloblastoma | 20 | 3.45 | 0 | 0.00 | 76 | 13.13 | 0 | 0.00 | 579 |
| PNET/Pineal | 1 | 0.37 | 0 | 0.00 | 19 | 7.12 | 0 | 0.00 | 267 |
| ATRT | 1 | 2.33 | 0 | 0.00 | 1 | 2.33 | 0 | 0.00 | 43 |
| Germ cell tumor | 4 | 3.70 | 0 | 0.00 | 18 | 16.67 | 0 | 0.00 | 108 |
| Teratoma | 0 | 0.00 | 0 | 0.00 | 0 | 0.00 | 0 | 0.00 | 13 |
| Other/unspecified CNS | 0 | 0.00 | 0 | 0.00 | 6 | 8.33 | 0 | 0.00 | 72 |
| Leukemia | 7 | 0.55 | 16 | 1.26 | 0 | 0.00 | 0 | 0.00 | 1,274 |

### Incidence

RIG incidence data were evaluated in two ways. We first investigated incidence of new RIG cases by year of original diagnosis, as a proportion of the total cases of new-onset first primary malignancies diagnosed in a given year that later went on to develop RIG. From 1975-2016, mean incidence per year in each cohort was as follows: Cohort 1 overall = 0.77% (range: 0-2.65%); Cohort 1a = 0.57% (range: 0-2.65%), Cohort 1b = 0.20% (0-1.37%), Cohort 2a = 1.47% (0-3.57%), and Cohort 2b = 1.92% (0-6.67%). For both Cohorts 1a and 1b, a trend toward decreasing incidence over time was observed via the 5-year moving average, and this was replicated when Cohort 1 was analyzed overall. The second method of evaluating RIG incidence involved determining the proportion of all CNS tumors diagnosed in a given year that were classified as RIG. From 1977-2016, the mean annual incidence of RIG using the Cohort 1a definition was 0.034% (range: 0-0.116%), while for Cohort 1b this was 0.011% (range: 0-0.057%). Combined, the mean annual incidence of RIG using the Cohort 1 definition was 0.04% (range: 0-0.17%). Using the 5-year moving average, a trend of increasing incidence over time was observed for Cohorts 1a, 1b, and Cohort 1 overall.

#### Latency to RIG diagnosis

Latency period between original diagnosis and development of RIG is shown in Table 4. Overall, median latency until RIG development for Cohort 1 was 11.1 years (minimum = 3.58 years, maximum = 34.42 years; Figure 1). Cohort 1a had a significantly longer median latency to RIG diagnosis as compared with Cohort 1b (12.0 years vs. 10.0 years, p=0.018; Figure 2).

**Figure 1.**
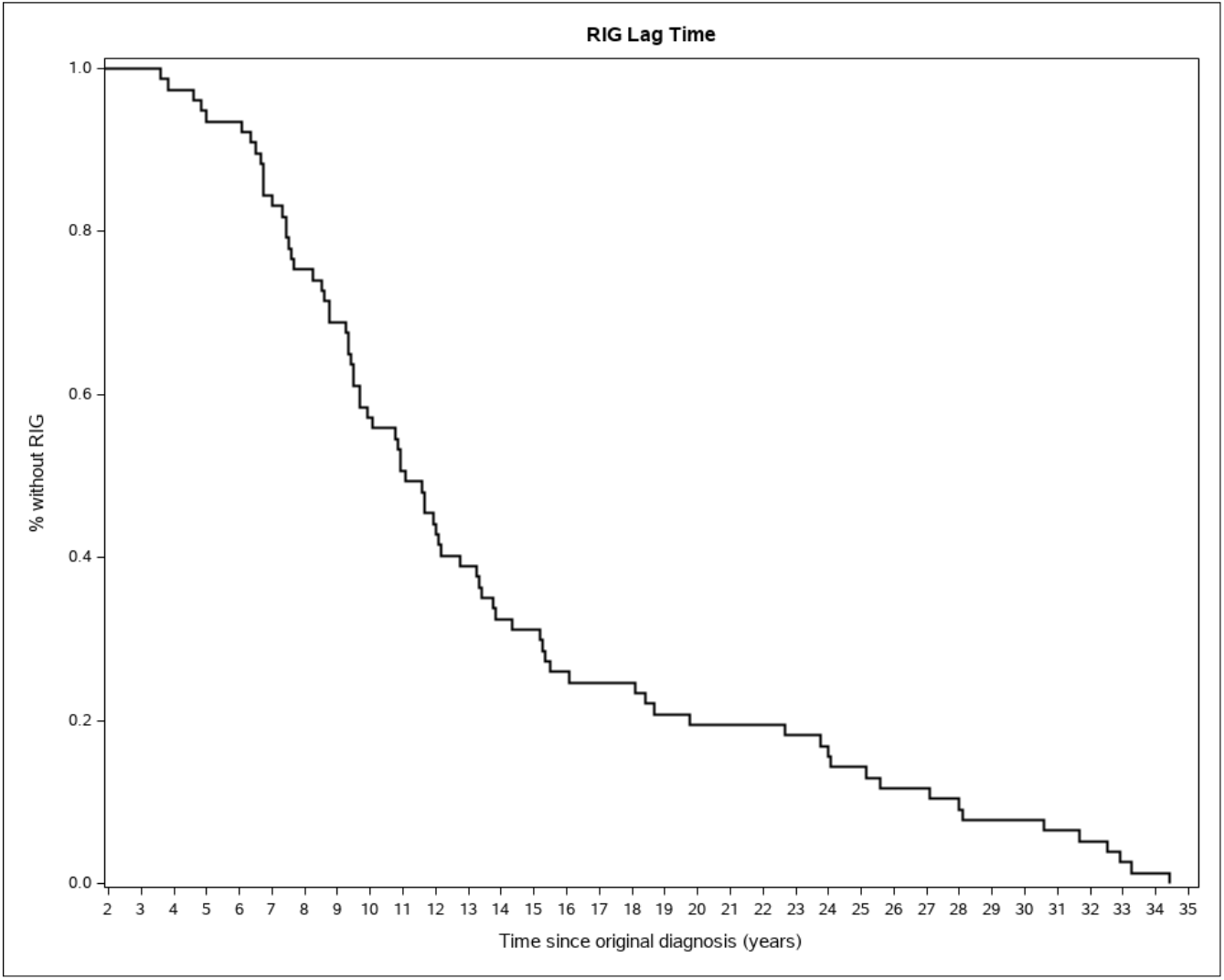
Lag time to RIG development, Cohort 1.

**Figure 2.**
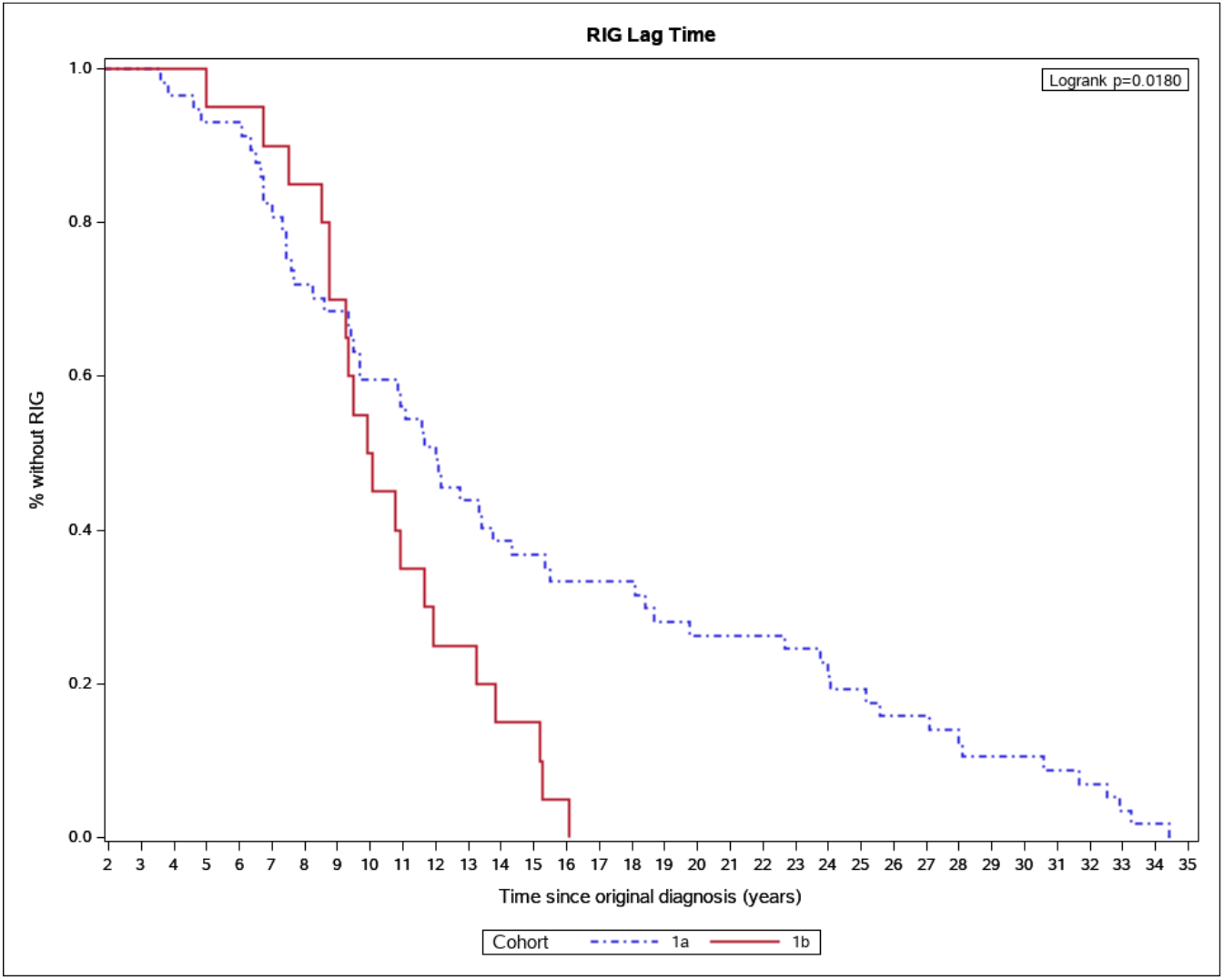
Lag time to RIG development, Cohort 1a vs. 1b.

#### Overall Survival

OS for Cohort 1 is displayed in Figure 3. Median OS for patients in Cohort 1 was 9.0 months. One year post-RIG diagnosis, median survival was 44.5% (95% CI = 32.8-55.5%). Two-year median OS was 15.9% (95% CI = 8.4-25.7%), and three-year median OS was 6.4% (95% CI = 2.1-14.1%). Over the course of the study period, 88% (50/57 patients) of patients in Cohort 1a died from RIG, while the mortality rate for patients in Cohort 1b was 80% (16/20 patients). In contrast, 43% (4,626/10,687) of control patients died over the course of the study period. Table 5 provides detailed information regarding overall deaths by cohort and total deaths broken down by original diagnosis.

**Figure 3.**
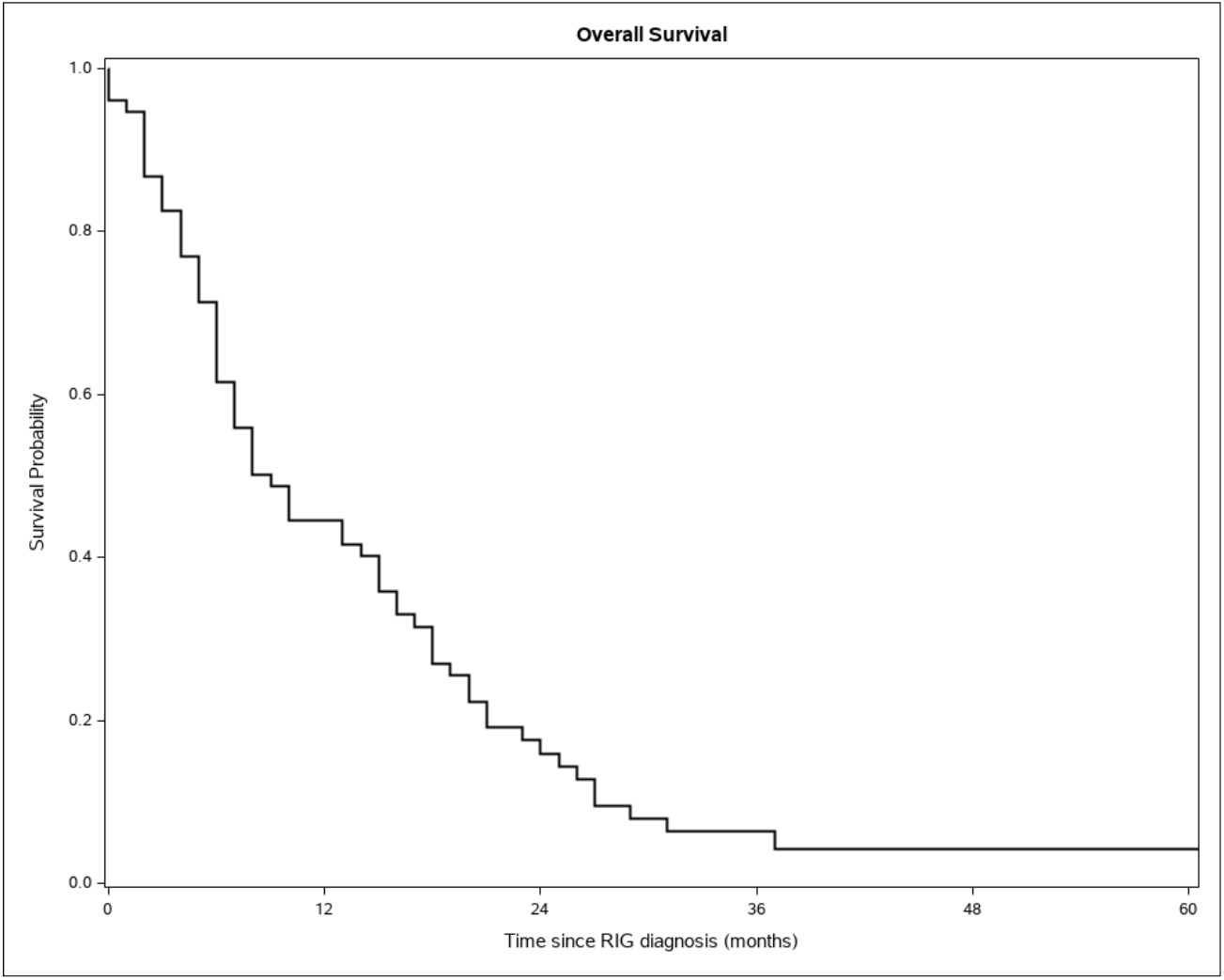
OS, Cohort 1.

We also compared median OS between patients by treatment type received for RIG. When comparing the group who received surgery to those in the No/Unknown group (Figure 4), no significant difference was observed in median OS (10.0 vs. 9.0 months; p=0.109). The group who received chemotherapy for RIG (Figure 5) had a median OS of 13.0 months, which was not significantly different from the No/Unknown Chemotherapy group who had a median OS of 6.0 months (p=0.174). Similarly, patients treated with radiation for RIG (Figure 6) had a median OS of 13.0 months, which was not significantly longer than the No/Unknown Radiation group with median OS of 6.0 months (p=0.133).

**Figure 4.**
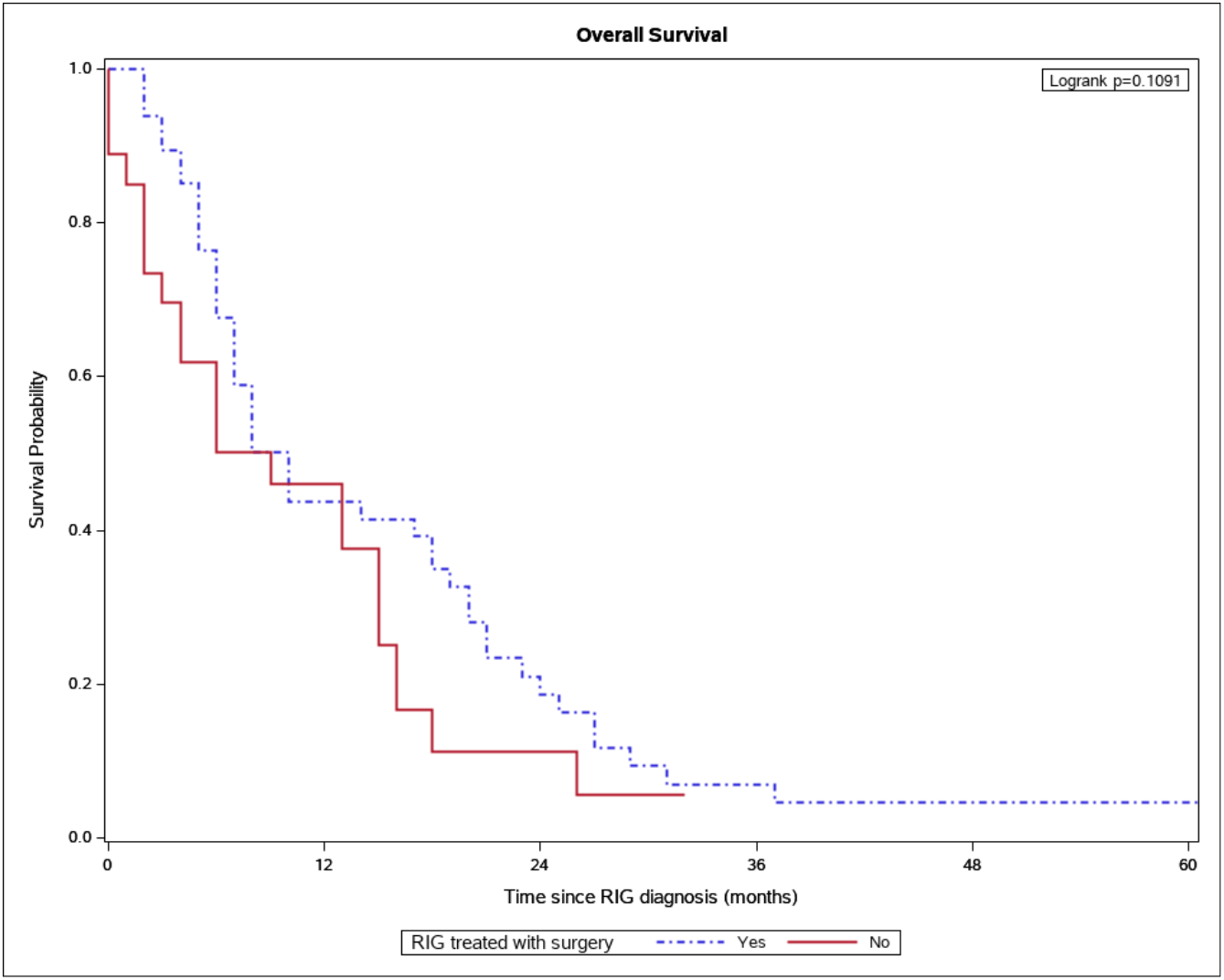
Median OS with and without surgery for RIG, Cohort 1.

**Figure 5.**
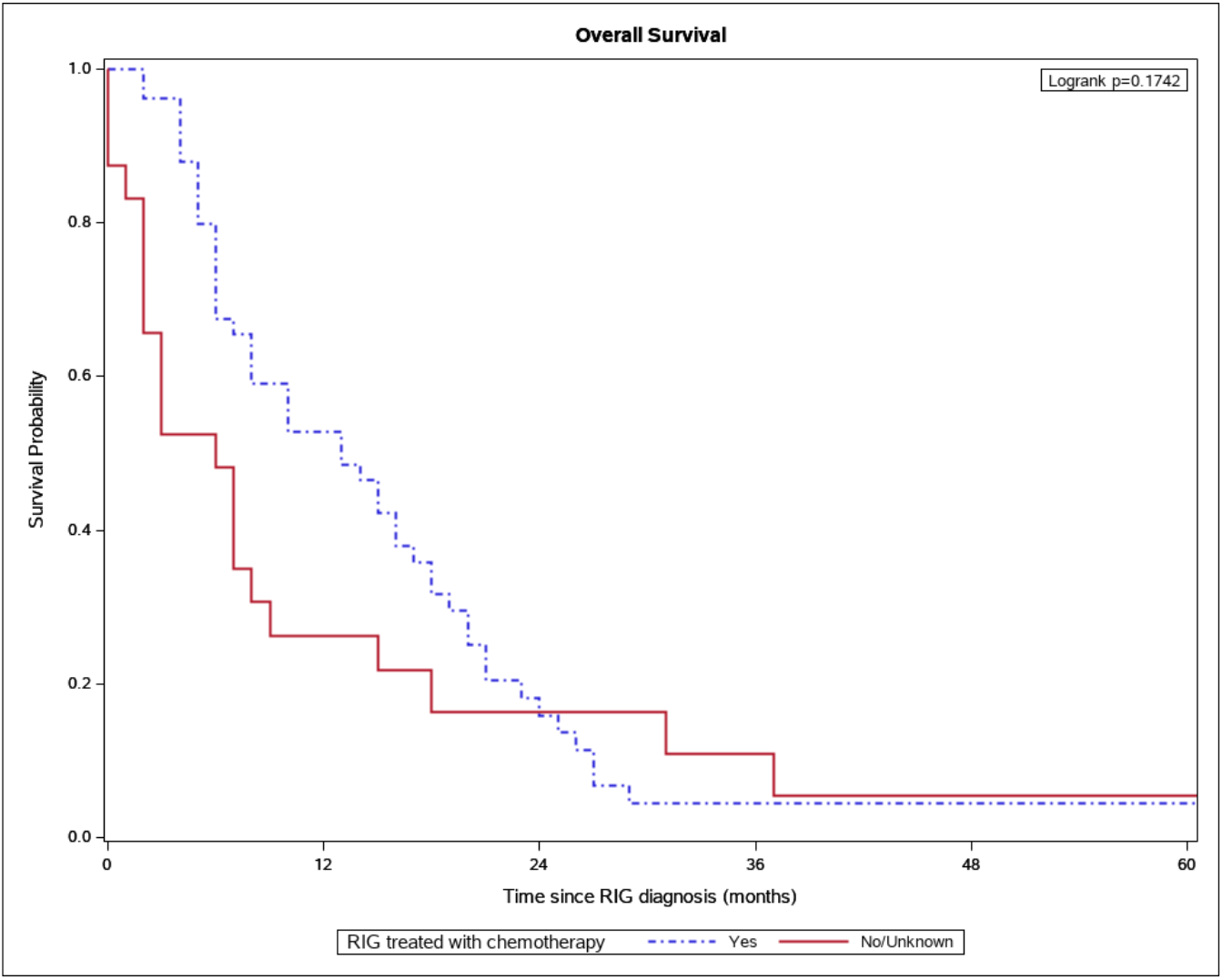
Median OS with and without chemotherapy for RIG, Cohort 1.

**Figure 6.**
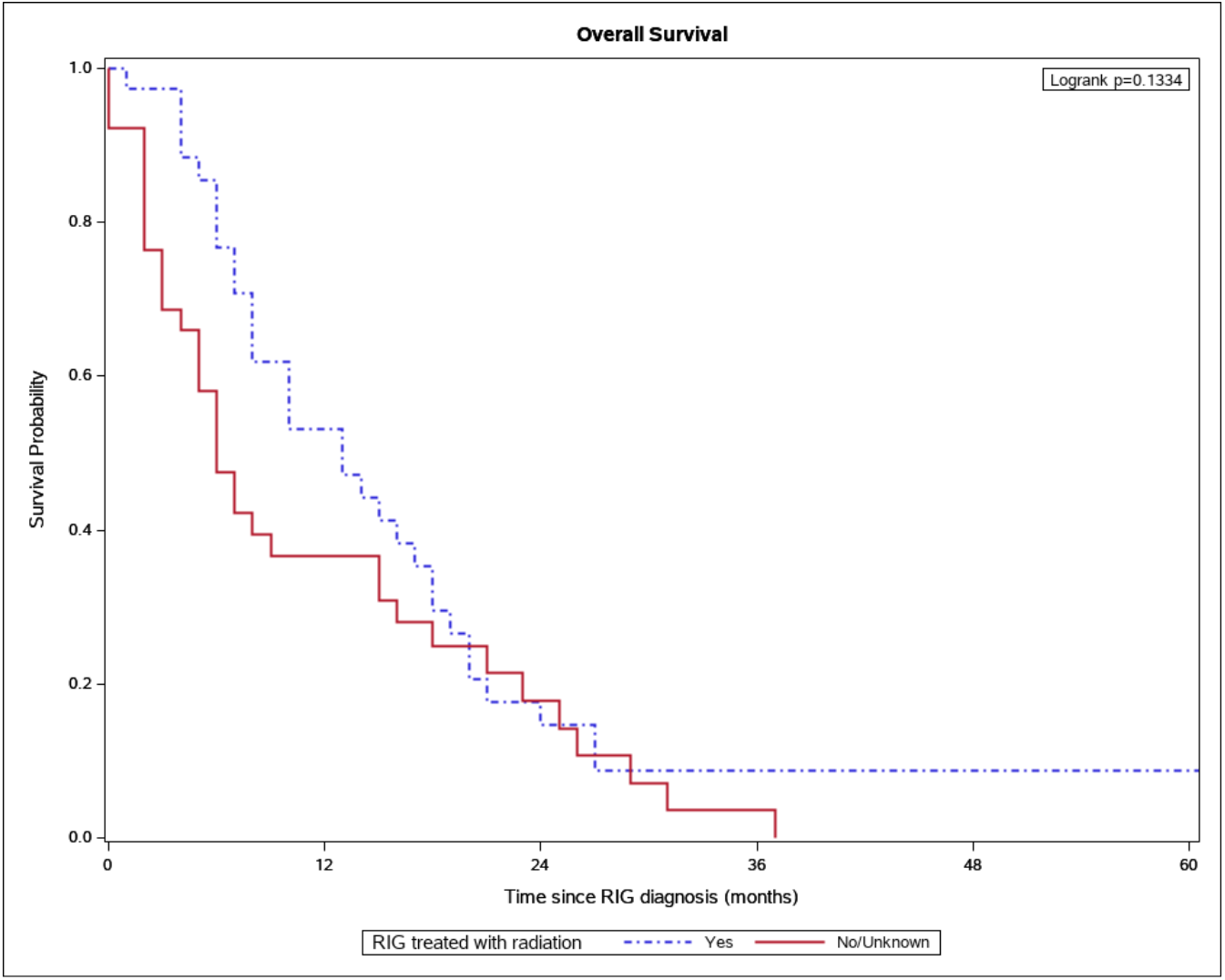
Median OS with and without radiation for RIG, Cohort 1.

## Discussion

Very little is understood about the epidemiology of RIG. The present study sought to characterize RIG using a population-based sample, including true incidence rates and changes in incidence over time, risk factors for RIG development, a timeline of RIG development following external-beam RT, and response to treatment with various modalities. We also aimed to better understand median OS for RIG. Our work expands upon prior literature by showing that RIG occurred in a small but substantial proportion of those who underwent treatment for pediatric tumors affecting the CNS, with a mean incidence rate of 0.77% in Cohort 1 by year of original diagnosis, and a mean incidence rate of 0.04% in Cohort 1 when analyzed by the proportion of all CNS tumors diagnosed in a given year that were classified as RIG. We demonstrated that RIG may develop far beyond the RT treatment period, with median lag time to RIG for Cohort 1 of 11.1 years and a range that extended more than 34 years beyond external-beam RT exposure. RIG appears to be a highly lethal malignancy with a dismal prognosis, with median OS for Cohort 1 of 9.0 months and only 6.4% of patients still living three years post-diagnosis.

Additionally, we identified a group of patients comprising Cohort 2 with possible undiagnosed RIG whose deaths occurred more than seven years after their original diagnosis, providing a rationale for more in-depth pathologic analysis of these patients’ tumors for determination of RIG status and treating these tumors accordingly.

Pediatric patients of all ages and races, male and female sex, and Hispanic and Non-Hispanic ethnicities were observed to develop RIG in our sample. The age of patients in Cohort 1b was significantly different from Controls, driven by the majority of Cohort 1b developing leukemias between ages 1-4 years. Age in Cohort 2b was also significantly different compared to Controls, with children ages 15-19 being more likely to develop glioma as their original primary malignancy. Notably, there were significantly fewer Hispanic patients in Cohorts 1a and 2b versus Controls, suggesting that Hispanic patients may be less likely to develop RIG post-RT. The most common original tumor diagnoses in confirmed RIG cases (Cohort 1) were leukemias, followed by medulloblastomas and gliomas. Though these findings may be specific to our analysis and our methodology used to collect data within SEER, it should be noted that these primary tumor types have previously been implicated as being among the most likely to go on to develop RIG.^22^

Our work was conducted using data derived from the SEER-18 Registries Custom Data, covering the years 1975-2016. SEER contains detailed information regarding patient demographics, cancer diagnoses, and survival over time; however, some patient-level information was less reliably included, including specific treatments received. With RIG being defined by exposure to RT, this made it difficult to precisely determine the incidence of RIG as some patients, especially those treated with cranial RT for leukemia, seemed to lack definitive data regarding whether they underwent external-beam RT. This necessitated further dividing Cohort 1 into Cohort 1a, comprised of those who were confirmed to have been treated with beam radiation for their first malignancy, and Cohort 1b, which included patients with leukemia whose RT treatment status was unknown. While we determined that mean incidence of RIG by year of original diagnosis for Cohort 1 was 0.77%, it is possible that this number could be closer to the mean incidence of 0.57% for Cohort 1a if not all patients with leukemia underwent RT.

Conversely, patients in Cohort 2 represented cases of possible undiagnosed RIG, as they died more than seven years following their first primary CNS malignancy diagnosis and treatment with external-beam RT. With primary CNS malignancies rarely recurring this late after initial treatment, it may be that at least some of these cases were not biopsied or were pathologically misclassified and would have fit molecular criteria for RIG. As we are unable to obtain samples and perform central pathological review for possible RIG reclassification, it cannot be definitively confirmed if any of these cases were indeed RIG. In the context of having no perfect registry available for data derivation, we included these patients to ensure that RIG cases were not missed and to highlight that the incidence numbers may be higher than reported here. These limitations within Cohorts 1 and 2 found within SEER raise the importance of establishing a RIG-specific registry which would contain detailed information about these patients, their disease courses, and the pathology of their tumors, providing investigators with a more robust database for further studies. Our group intends to establish such a RIG registry moving forward for the purpose of better understanding this disease.

Mean incidence of RIG by year of original diagnosis generally appeared to decline over time between 1975-2016. As we observed that median latency to RIG was 11.1 years after diagnosis of first malignancy, it is likely that there has not yet been sufficient lag time for patients diagnosed and treated with RT in more recent years to go on to develop RIG, which is likely driving the appearance of decreased case rates. Meanwhile, when analyzing mean RIG incidence as a proportion of all CNS tumors diagnosed at any age in a given year, we found that incidence appeared to increase over time. As incidence rates of pediatric brain tumors have been increasing with time,^23^ this may be a product of a growing number of children receiving cranial RT. The threat of RIG as a sequela of RT may be growing, warranting further investigation of potential therapies useful in treating it.

The potential for rising incidence rates of RIG raises the importance of understanding its impact on patients’ lives. Our observed median OS of 9.0 months for patients with RIG portends a dismal prognosis for these individuals and is comparable to that of diffuse intrinsic pontine glioma, a pHGG with arguably the worst prognosis and which has proven to be exceedingly difficult to treat.^24,25^ Three years following diagnosis of RIG, only 6.4% of patients remained alive in our sample. These figures, while jarring, make sense in the context of our finding that no currently available treatments including surgery, RT, or chemotherapy appeared to significantly improve median OS. While this contradicts a prior report showing benefit of resection for RIG over other modalities,^15^ this study’s findings may have been influenced by a very small sample size. Our findings are consistent with literature showing that RIG is poorly responsive to treatment,^13^ and highlight the need for development of better preclinical models and the initiation of more clinical trials for therapies specifically targeting RIG.

Our work is largely concordant with the small but existing body of RIG literature. Incidence of RIG development after cranial RT has been estimated to occur in ∼0.5-3% of patients^13,26^ after a median latency period of 9-15 years,^27,28^ with a median OS of 9-11 months and 2-year survival rate of approximately 20%.^22,27^ While these retrospective cohort and comprehensive review-style studies provide an important foundation for RIG epidemiology, we expand upon these here by providing annual incidence rates over four decades both by year of original diagnosis and as a proportion of all CNS tumors diagnosed in a given year. This study is also the first of its kind to use a large, population-based SEER dataset in an attempt to better characterize incidence and impact of RIG with greater population representation than prior studies. A notable finding was that RIG development occurred as late as ∼34 years after original diagnosis in our sample, indicating the need for clinicians to be aware of the potential for RIG to occur many years after RT exposure. This may have implications for long-term follow-up for patients treated with cranial RT. It is also interesting to note the significantly shorter median latency to RIG for Cohort 1b compared to Cohort 1a in our sample, which may be attributable to patients with leukemias receiving whole-brain radiation for CNS prophylaxis in the earlier years of the study period.

There are several limitations to our work. Using data from SEER registries spanning 1975-2016, only 9% of the population was represented within registry data from 1975-1991, while approximately 28% of the population was covered between 2000-2016. This may limit the generalizability of these findings to the larger U.S. population outside of geographic areas covered within these registries. Given that our data span several decades, patients who were diagnosed more recently had shorter follow-up and thus may misleadingly reflect a lower incidence of RIG. Inherent to the SEER database, it is possible that patient demographics such as race and ethnicity were misclassified, and a substantial number of patients had limited information regarding treatment history. We also opted to include only patients who received external-beam RT, which excluded patients who may have developed RIG as a result of other radiation treatment techniques and may underestimate the overall incidence of RIG.

## Conclusion

Using a large, population-based sample of pediatric patients with tumors affecting the CNS within the SEER registries, we characterized the annual incidence of RIG by year of original diagnosis and as a proportion of all CNS tumors diagnosed at any age in a given year between 1975-2016. We found incidence rates concordant with existing literature and observed median latency to RIG that was beyond the previously established understanding of the timeframe within which RIG usually occurs. This suggests that clinicians should be aware of the potential need for following patients who underwent cranial RT as children beyond the first 15 years after treatment. We also found that RIG carries a very poor prognosis, with median OS of 9.0 months and no particularly effective treatment options currently available. Focused effort should be given toward developing better preclinical models of RIG and conducting translational and clinical studies of therapies specifically targeted at this treatment-resistant tumor subtype. Establishment of a national registry of RIG patients and tumor pathology samples would be an important first step toward this effort.

## Supporting information

COI Statement

STROBE Checklist Case-Control

## Data Availability

All data produced in the present study are available upon reasonable request to the authors.

## References

1. Ostrom QT, de Blank PM, Kruchko C, Petersen CM, Liao P, Finlay JL, Stearns DS, Wolff JE, Wolinsky Y, Letterio JJ et al. (2015) Alex’s lemonade stand foundation infant and childhood primary brain and central nervous system tumors diagnosed in the United States in 2007–2011. Neuro Oncol 16 (Suppl 10), x1– x36.

2. Finlay JL, Boyett JM, Yates AJ, Wisoff JH, Milstein JM, Geyer JR, Bertolone SJ, McGuire P, Cherlow JM & Tefft M (1995) Randomized phase III trial in childhood high-grade astrocytoma comparing vincristine, lomustine, and prednisone with the eightdrugs-in-1-day regimen. Childrens Cancer Group. J Clin Oncol 13, 112–123.

3. Sposto R, Ertel IJ, Jenkin RD, Boesel CP, Venes JL, Ortega JA, Evans AE, Wara W & Hammond D (1989) The effectiveness of chemotherapy for treatment of high grade astrocytoma in children: results of a randomized trial. A report from the Childrens Cancer Study Group. J Neurooncol 7, 165–177.

4. Lulla RR, Saratsis AM, Hashizume R. Mutations in chromatin machinery and pediatric high-grade glioma. Sci Adv (2016) 2(3):e1501354. 10.1126/sciadv.1501354

5. Meel MH, Schaper SA, Kaspers GJL, Hulleman E. Signaling pathways and mesenchymal transition in pediatric high-grade glioma. Cell Mol Life Sci (2018) 75(5):871–87. 10.1007/s00018-017-2714-7

6. Wierzbicki K, Ravi K, Franson A, Bruzek A, Cantor E, Harris M, et al. Targeting and Therapeutic Monitoring of H3K27M-Mutant Glioma. Curr Oncol Rep (2020) 22(2):19. 10.1007/s11912-020-0877-0

7. DeNunzio NJ, Yock TI. Modern Radiotherapy for Pediatric Brain Tumors. Cancers (Basel). 2020;12(6):1533. doi:10.3390/cancers12061533.

8. Pui CH, Mullighan CG, Evans WE, Relling MV. Pediatric acute lymphoblastic leukemia: where are we going and how do we get there? Blood. 2012;120:1165–74.

9. Sturm D, Pfister SM, Jones DTW. Pediatric Gliomas: Current Concepts on Diagnosis, Biology, and Clinical Management. J Clin Oncol (2017) 35(21):2370–7. 10.1200/JCO.2017.73.0242

10. Sharda N, Yang C-R, Kinsella T, Boothman D. Radiation Resistance. In: Bertino JR, editor. Encyclopedia of Cancer, 2nd ed. New York: Academic Press; (2002). p. 1–11.

11. Metselaar DS, du Chatinier A, Stuiver I, Kaspers GJL, Hulleman E. Radiosensitization in Pediatric High-Grade Glioma: Targets, Resistance and Developments. Front Oncol. 2021;11:662209. Published 2021 Apr 1. doi:10.3389/fonc.2021.662209

12. Donson AM, Erwin NS, Kleinschmidt-DeMasters BK, Madden JR, Addo-Yobo SO & Foreman NK (2007) Unique molecular characteristics of radiation-induced glioblastoma. J Neuropathol Exp Neurol 66, 740–749.

13. DeSisto J, Lucas JT Jr, Xu K, Donson A, Lin T, Sanford B, et al. Comprehensive molecular characterization of pediatric radiation-induced high-grade glioma. Nat Commun. 2021 Sep 20;12(1):5531. doi: 10.1038/s41467-021-25709-x.

14. Armstrong GT, Liu Q, Yasui Y, Neglia JP, Leisenring W, Robison LL & Mertens AC (2009) Late mortality among 5-year survivors of childhood cancer: a summary from the Childhood Cancer Survivor Study. J Clin Oncol 27, 2328–2338.

15. Carret AS, Tabori U, Crooks B, Hukin J, Odame I, Johnston DL, Keene DL, Freeman C, Bouffet E & Canadian Pediatric Brain Tumour Consortium (2006) Outcome of secondary high-grade glioma in children previously treated for a malignant condition: a study of the Canadian Pediatric Brain Tumour Consortium. Radiother Oncol 81, 33–38.

16. Chatwin HV, Cruz J, Green AL. Pediatric high-grade glioma: moving toward subtype-specific multimodal therapy. FEBS J. 2021 Feb 1. doi: 10.1111/febs.15739. Epub ahead of print. PMID: 33523591.

17. Deng MY, Sturm D, Pfaff E, Sill M, Stichel D, Balasubramanian GP, et al. Radiation-induced gliomas represent H3-/IDH-wild type pediatric gliomas with recurrent PDGFRA amplification and loss of CDKN2A/B. Nat Commun. 2021 Sep 20;12(1):5530. doi: 10.1038/s41467-021-25708-y.

18. Lopez GY, Van Ziffle J, Onodera C, Grenert JP, Yeh I, Bastian BC, Clarke J, Oberheim Bush NA, Taylor J, Chang S et al. (2019) The genetic landscape of gliomas arising after therapeutic radiation. Acta Neuropathol 137, 139–150.

19. Surveillance, Epidemiology, and End Results (SEER) Program (www.seer.cancer.gov) SEER*Stat Database: Incidence - SEER 18 Regs Custom Data (with additional treatment fields), Nov 2018 Sub (1975-2016 varying) - Linked To County Attributes - Total U.S., 1969-2017 Counties, National Cancer Institute, DCCPS, Surveillance Research Program, released April 2019, based on the November 2018 submission.

20. Hiraki T, Fukuoka K, Mori M, et al. Application of genome-wide DNA methylation analysis to differentiate a case of radiation-induced glioblastoma from late-relapsed medulloblastoma. J Neuropathol Exp Neurol. 2021; 80(6):552–557.

21. Yang SY, Wang KC, Cho BK, et al. Radiation-induced cerebellar glioblastoma at the site of a treated medulloblastoma: case report. J Neurosurg. 2005; 102(4 Suppl):417–422.

22. Yamanaka R, Hayano A, and Kanayama T. Radiation-induced gliomas: a comprehensive review and meta-analysis. Neurosurg Rev. 2018; 41:719–731.

23. Howlader NN, Noone AM, Krapcho ME, Miller D, Brest A, Yu ME, Ruhl J, Tatalovich Z, Mariotto A, Lewis DR, Chen HS. SEER cancer statistics review, 1975–2016. National Cancer Institute. 2019 Apr;1.

24. Hargrave D, Bartels U, Bouffet E. Diffuse brainstem glioma in children: critical review of clinical trials. Lancet Oncol. 2006 Mar;7(3):241–8. doi: 10.1016/S1470-2045(06)70615-5. PMID: 16510333.

25. Jansen MH, van Vuurden DG, Vandertop WP, Kaspers GJ. Diffuse intrinsic pontine gliomas: a systematic update on clinical trials and biology. Cancer Treat Rev. 2012 Feb;38(1):27–35. doi: 10.1016/j.ctrv.2011.06.007. Epub 2011 Jul 20. PMID: 21764221.

26. Aherne NJ, Murphy BM. Radiation-Induced Gliomas. Crit Rev Oncog. 2018;23(1-2):113-118. doi: 10.1615/CritRevOncog.2018025740. PMID: 29953370.

27. Elsamadicy AA, Babu R, Kirkpatrick JP, and Adamson DC. Radiation-induced malignant gliomas: A current review. World Neurosurg. 2015; 83(4):530–542.

28. Prasad G, Haas-Kogan DA. Radiation-induced gliomas. Expert Rev Neurother. 2009;9(10):1511–1517. doi:10.1002/pmic.200800802

